# Association of pitavastatin use with bone markers and bone mineral density in postmenopausal women

**DOI:** 10.1101/2024.08.14.24312019

**Authors:** Jihye Hyun, Minji Sohn, Hyeran Oh, Soo Lim

## Abstract

**BACKGROUND:** The study aimed to evaluate the effects of pitavastatin therapy on biochemical markers of bone turnover and bone mineral density (BMD) in postmenopausal women with osteopenia or osteoporosis and hypercholesterolemia.

**METHODS AND RESULTS:** This prospective observational study recruited 70 postmenopausal Korean women who were administered pitavastatin. Changes in BMD at the femoral neck (FN), total hip (TH), and lumbar spines (LS), as well as laboratory values related to bone turnover markers and lipid profiles, including lipoprotein subfractions, were assessed over 12 months. BMD was further observed during regular clinic visits up to 4 years. A total of 67 patients completed 12 months of pitavastatin therapy. There were no significant changes in BMD (–0.001 ± 0.026 at FN, –0.005 ± 0.032 at TH, and 0.002 ± 0.037 at LS; all Ps > 0.05). Bone markers did not change except for procollagen type 1 N-propeptide (P1NP), which slightly decreased (–4.0 ± 10.3, *P* < 0.05). Low-density lipoprotein (LDL) cholesterol levels decreased by 36% to 39% from baseline, and the LDL subfraction score decreased from 2.85 ± 1.79 to 1.80 ± 0.63 (*P* < 0.001). During the extended observation period, BMD at FN and TH decreased by 0.85% and 1.69% per year, respectively, while BMD at the LS was preserved.

**CONCLUSIONS:** In postmenopausal women with osteopenia or osteoporosis requiring lipid-lowering therapy, one year of pitavastatin maintained BMD and effectively controlled cholesterol levels. BMD changes during statin therapy were more favorable than the natural decline. These findings suggest pitavastatin can be safely used in menopausal women without compromising bone health.

## Introduction

Statins, primarily used to lower cholesterol levels and prevent cardiovascular diseases, have been investigated for their additional effects on bone health.^1^ A statin was associated with a lower risk of fractures in the elderly from a UK national database,^2^ while other studies reported conflicting results, showing no effect on fractures in postmenopausal women or even worsening bone mineral density (BMD) in type 2 diabetes.^3,4^ These indicate that statins needs to be used cautiously and tailored to specific cases based on the type of statins. Given the growing prevalence of osteoporosis in aging populations who might also require statins, the relationship between statins and bone health needs further evidence.

Osteoporosis is a skeletal disorder characterized by weakened bone strength and is increasingly prevalent in rapidly aging societies.^5^ Traditional risk factors for osteoporosis include aging, hormonal changes, inadequate dietary calcium and vitamin D, and some medications.^6^ The major determinant of bone loss in women during middle age is menopause,^7^ but concomitant long-term medication may affect their bone health.

A systematic review and meta-analysis of 33 clinical trials found that statin treatment was associated with increased BMD at the total hip (TH) and lumbar spines (LS), and improved the bone formation marker, such as osteocalcin, while having a neutral effect on BMD at the femoral neck (FN), bone-specific alkaline phosphatase, and C-terminal telopeptide (CTX).^8^ Most of the findings were from observational studies, with limited randomized controlled trials showing a neutral effect on BMD.

Pitavastatin has moderate-intensity cholesterol-lowering effects^9^ and a long duration of action, providing convenience to patients regardless of the time of administration.^10^ With minimal metabolism through the cytochrome P450 enzymes, pitavastatin is primarily metabolized through UDP-glucuronosyltransferase enzymes and has reduced potential for drug interaction.^10,11^ These characteristics might be beneficial for the elderly who might use multiple medications from the aspect of drug-drug interaction.

In this study, we aimed to investigate the effects of pitavastatin therapy on bone health in postmenopausal women with osteopenia or osteoporosis and concurrent hypercholesterolemia. We expected to provide valuable insights into its potential effects on patients with osteopenia or osteoporosis and hypercholesterolemia.

## Patients and Methods

### Data Availability Statement

The datasets collected and investigated in this study are available from the corresponding author upon a reasonable request.

### Study Design and Setting

This study was designed as a prospective, single-arm, open label study and was approved by an independent ethics committee/Institutional Review Board (B-1902-520-004). It was registered at clinicaltrials.gov (NCT06359353) and conducted at Seoul National University Bundang Hospital. All participants provided written informed consent prior to participating in the study.

Our study size of 70 participants was set based on a reference study^12^ that reported a significant reduction in serum N-terminal telopeptide of type 1 collagen (NTX) from 15.53 ± 4.2 nM BCE/mM Cr to 12.06 ± 2.6 nM BCE/mM Cr after three months of pitavastatin treatment. We targeted 66 participants, accounting for a 10% dropout rate. We included postmenopausal women aged 75 years or younger. Eligibility was confirmed through initial screenings and medical records review, focusing on patients who were diagnosed with osteopenia or osteoporosis (–3.0 ≤ T-score ≤ –1.5), initiating pitavastatin treatment for hypercholesterolemia for the first time, and presenting with baseline bone resorption markers (CTX ≥ 0.300 ng/mL or urinary NTX > 16.5 nM BCE/mM Cr).

Patients were excluded based on the following criteria: (1) history of statin use for more than one month within the three months prior to the study; (2) treatment with oral or injectable glucocorticoids for over one week in the same period; (3) current use of thiazolidinediones; (4) ongoing treatment for malignant tumors.

All participants received pitavastatin orally at a daily dosage of 4 mg for 52 weeks or more. While most patients maintained the dosage of 4 mg of pitavastatin once daily throughout the study period, the dosage was decreased to 2 mg daily at physician’s discretion.

### Variables and Outcome Measurements

The primary outcome was the change in BMD after 12 months of pitavastatin therapy. Secondary outcomes included the changes in osteocalcin, procollagen type 1 N-terminal propeptide (P1NP), CTX, and NTX after 6 and 12 months. Additionally, laboratory values related to patients’ general status, including cholesterol levels, glycated haemoglobin (HbA1c), liver and kidney function, as well as those related to bone mineral metabolism such as 25-hydroxyl-vitamin D (25OH-vitamin D) and parathyroid hormone (PTH) levels, were assessed at 3, 6 and 12 months.

BMD was measured using dual-energy X-ray absorptiometry (DXA) with a QDR 4500 device (Hologic, Waltham, MA, USA). We also used the T-score, which compares an individual’s BMD with the mean value for young, normal women and expresses the differences as a standard deviation score.^13^ Changes in BMD and T-scores from baseline to 12 months post-treatment at the following sites: left hip (including FN, TH), and LS (L1, L2, L3, L4, and the average from L1 to L4) were observed. BMD was further investigated as part of an extended protocol in year 2, and also collected if examined during regular clinic visits for up to 4 years.

Body composition including body weight, height, and body mass index (BMI), was measured by standard methods with the participants wearing light clothing.

Biochemical markers of bone metabolism were measured: Osteocalcin and P1NP levels using the Elecsys N-MID Osteocalcin and Total P1NP assays (Roche), respectively. Urine NTX and CTX levels were assessed via an NTX ELISA kit (Inverness Medical) and the Elecsys B-CrossLaps assay (Roche). Serum calcium, phosphorus, 25OH-vitamin D, and PTH were evaluated using colorimetry (Hitachi 747), HPLC (Variant II Turbo, Bio-Rad), and chemiluminescence (LIAISON; DiaSorin), respectively.

Plasma glucose and HbA1c were determined using the glucose oxidase method (Hitachi 747) and HPLC (Variant II Turbo). Serum AST, ALT, creatinine, and eGFR were measured with the Architect Ci8200 (Abbott). Cholesterol profiles were analyzed with Hitachi’s Clinical Chemistry Analyzer. Urinary albumin and creatinine were quantified by turbidimetry (502X, A&T) and the Jaffe method (Hitachi 7170).

Lipoprotein subfractions were also measured using the Quantimetrix LDL Lipoprint System (Quantimetrix Corporation, Redondo Beach, CA, USA) following the manufacturer’s instructions.^14^ This method used high-resolution polyacrylamide gel electrophoresis to separate and measure very low-density lipoprotein (VLDL), intermediate-density lipoprotein (IDL), and LDL subfractions. IDL was divided into large (C), medium (B), and small (A) subfractions. The LDL subfraction score was calculated using the weighted area under the curve (AUC) of LDLs.

### Statistical Analysis

All patients in the study were enrolled in the analysis. Clinical characteristics are presented as means ± standard deviation (SD). To compare baseline and post-treatment values across all successive pairs during the follow-up period for each patient, we used paired *t*-tests to analyze changes in clinical variables. The mean annual percent change in BMD was calculated by dividing the difference between the baseline BMD and follow-up measurements of BMD by the baseline BMD, then dividing by the study period (in years) and multiplying by 100. Additionally, we analyzed over 4 years (1,614 days) of observations to compare the fitted slope of BMD changes in our cohort with those reported in other long-term studies (4 years or more) on postmenopausal women who had no history of treatments affecting bone health, aiming to compare with natural BMD changes. A *t*-test was conducted to compare our estimated slope, derived from a simple linear regression model, with the average slopes of the international and Korean cohorts. For data management, participants who did not take their prescribed diabetes medication during the study period were excluded from the glucose level analysis. Also, BMD and T-score values that were deemed inappropriate due to specific reasons such as pin fixation, cement injection, etc., were excluded from the analysis, to account for variations in personal examination.

All statistical analyses were conducted using R software, version 4.3.3 (R Foundation for Statistical Computing, Vienna, Austria). A *P*-value of < 0.05 was considered statistically significant, indicating evidence of a meaningful difference.

## Results

### Baseline Characteristics of Study Participants

A total of 70 patients with an average age of 65.3 ± 5.3 years were enrolled (**Table S1**). The most common underlying diseases were diabetes (31.4%), osteopenia (30%), and hypothyroidism (22.9%). The lipid profiles of the study group showed a total cholesterol levels of 229 ± 35 mg/dL, triglyceride levels of 127 ± 54 mg/dL, and LDL-cholesterol levels of 145 ± 25 mg/dL. The serum 25OH-vitamin D levels averaged 26.2 ± 10.1 ng/mL.

### Changes in Lipid Profiles and Biochemical Parameters over 12 months

***Tables 1*** and ***2*** present the changes in lipid profiles and biochemical parameters in 67 patients who received 12 months of pitavastatin treatment. LDL-cholesterol levels decreased from 145.7 ± 24.1 mg/dL to 89.1 ± 20.2 mg/dL at 6 months and to 93.7 ± 30.1 mg/dL at 12 months, showing declines of 38.8% and 35.7%, respectively. Four people reduced their dosage from 4 mg to 2 mg (***Table 1***). Remnant cholesterol, triglycerides, VLDL, IDL-C, IDL-B, IDL-A, and LDL particles (LDL Large 1 to LDL Small 5) also decreased during the 1-year observation (*P* < 0.001 for all time points).

**Table 1.**
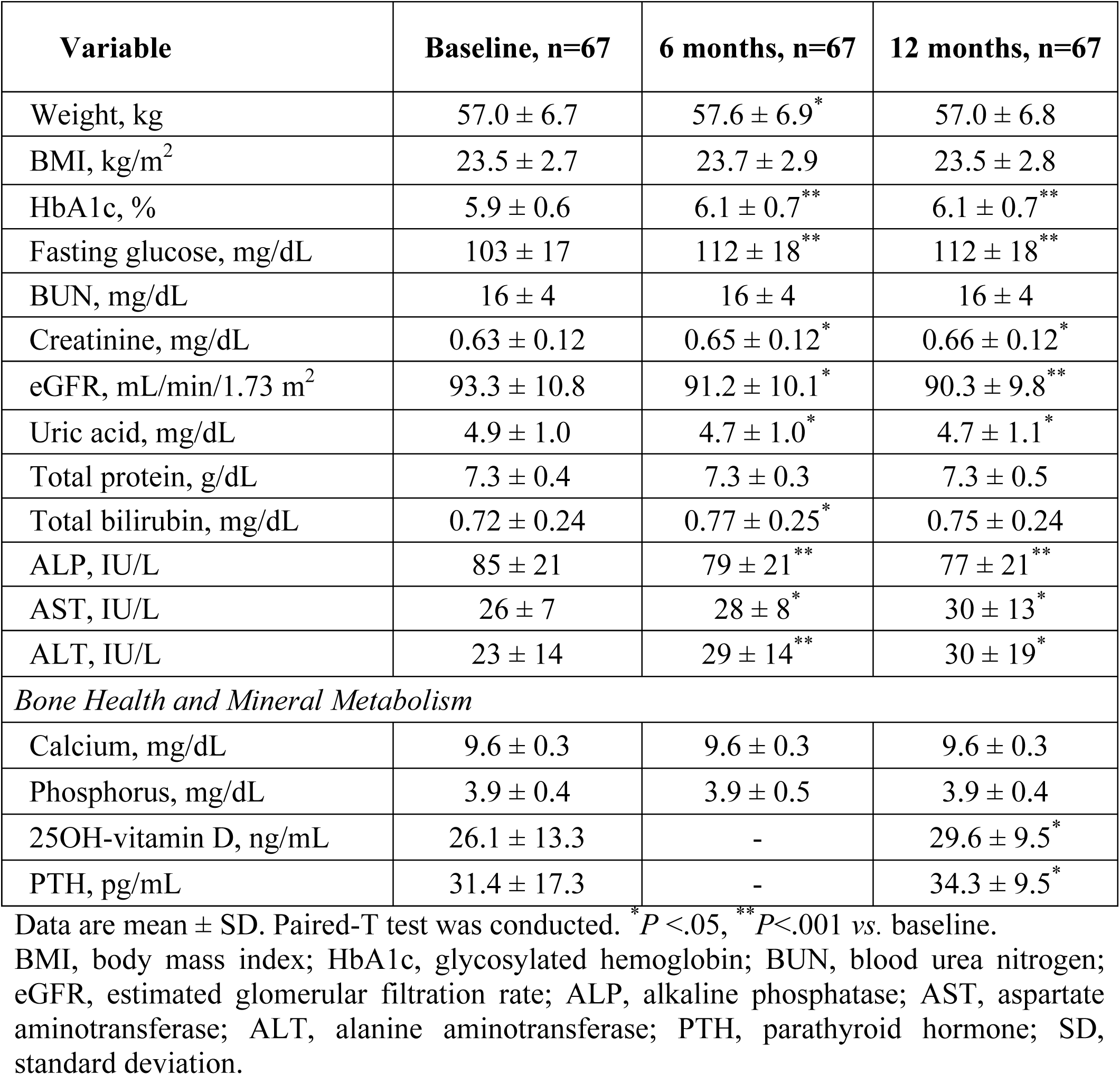
Patient characteristics over 12 months.

**Table 2.** Changes in lipid profiles over 12 months.

| Variable | Baseline, n=67 | 6 months, n=67 | 12 months, n=67 |
| --- | --- | --- | --- |
| Total cholesterol, mg/dL | 231.4 ± 35.7 | 162.7 ± 28.1<sup>**</sup> | 173.2 ± 38.0<sup>**</sup> |
| LDL-cholesterol, mg/dL | 146.1 ± 24.0 | 88.7 ± 19.9<sup>**</sup> | 94.0 ± 30.0<sup>**</sup> |
| HDL-cholesterol, mg/dL | 64.0 ± 17.4 | 64.1 ± 16.0 | 61.6 ± 16.8<sup>*</sup> |
| Triglycerides, mg/dL | 126.3 ± 54.2 | 104.5 ± 39.7<sup>**</sup> | 109.2 ± 42.5<sup>*</sup> |
| Non HDL-cholesterol, mg/dL | 167.4 ± 28.2 | 98.6 ± 21.4<sup>**</sup> | 111.6 ± 33.0<sup>**</sup> |
| Remnant cholesterol, mg/dL | 21.3 ± 7.2 | 9.9 ± 6.1<sup>**</sup> | 17.6 ± 10.2<sup>*</sup> |
| *Subpopulation analysis* |  |  |  |
| VLDL, mg/dL | 31.7 ± 11.3 | 23.2 ± 7.1<sup>**</sup> | 24.1 ± 8.0<sup>**</sup> |
| IDL-C, mg/dL | 24.5 ± 6.4 | 15.9 ± 4.1<sup>**</sup> | 16.4 ± 4.0<sup>**</sup> |
| IDL-B, mg/dL | 13.5 ± 3.9 | 9.3 ± 2.8<sup>**</sup> | 9.5 ± 3.2<sup>**</sup> |
| IDL-A, mg/dL | 15.1 ± 4.6 | 11.6 ± 3.8<sup>**</sup> | 12.4 ± 4.1<sup>**</sup> |
| LDL Large-1, mg/dL | 39.2 ± 15.7 | 28.6 ± 7.7<sup>**</sup> | 31.7 ± 10.4<sup>**</sup> |
| LDL Large-2, mg/dL | 32.7 ± 12.4 | 19.6 ± 6.8<sup>**</sup> | 23.5 ± 10.6<sup>**</sup> |
| LDL Small-3, mg/dL | 14.0 ± 7.8 | 4.9 ± 4.0<sup>**</sup> | 7.1 ± 7.6<sup>**</sup> |
| LDL Small-4, mg/dL | 6.2 ± 6.2 | 1.0 ± 2.8<sup>**</sup> | 1.4 ± 2.9<sup>**</sup> |
| LDL Small-5, mg/dL | 2.1 ± 4.2 | 0.3 ± 2.1<sup>*</sup> | 0.1 ± 0.5<sup>*</sup> |
| LDL Small-6, mg/dL | 0.3 ± 1.3 | 0.0 ± 0.1<sup>*</sup> | 0.0 ± 0.0 |
| LDL Small-7, mg/dL | 0.0 ± 0.3 | 0.0 ± 0.0 | 0.0 ± 0.0 |
| LDL subfraction score | 2.86 ± 1.78 | 1.79 ± 0.62<sup>**</sup> | 1.79 ± 0.50<sup>**</sup> |

Data are presented as mean ± standard deviation. Paired-T test was conducted. <sup>\*</sup>*P* <.05, <sup>\*\*</sup>*P* <.001 vs. baseline.
LDL, low-density lipoprotein; HDL, high-density lipoprotein; VLDL, very low-density lipoprotein; IDL, intermediate-density lipoprotein.

The 25OH-vitamin D and PTH levels increased from baseline to 12 months (*P* = 0.012 and *P* = 0.038, respectively) (***Table 1***), while calcium levels did not change over the 12 months.

### BMD and T-score Changes with Bone Turnover Markers over 12 months

BMD measurements at all sites—FN, TH, and LS—showed no significant changes over 12 months (***Table 3***): a –0.16% decrease in BMD at FN (absolute change: –0.001 ± 0.026), a – 0.66% decrease at TH (absolute change: –0.005 ± 0.032), and a 0.26% increase at LS (absolute change: 0.002 ± 0.037). Similarly, T-scores also did not show significant changes at any site. Although T-scores at the LS showed slight increases with absolute changes ranging from 0.03 to 0.08, these changes were not statistically significant.

**Table 3.** Changes in BMD and T-score comparisons between baseline and 12 months.

| Site of measurement | Baseline, n=67 | 12 months, n=67 | Baseline vs. 12 months |  |  |
| --- | --- | --- | --- | --- | --- |
|  |  |  | Absolute change | Mean percent change/year (%) | <sup>a</sup>P-value |
| *BMD, g/cm<sup>2</sup>* |  |  |  |  |  |
| Femoral Neck | 0.618 ± 0.057 | 0.617 ± 0.061 | −0.001 ± 0.026 | −0.16 | 0.797 |
| Total Hip | 0.759 ± 0.063 | 0.753 ± 0.067 | −0.005 ± 0.032 | −0.66 | 0.201 |
| Lumbar Spine | 0.784 ± 0.068 | 0.786 ± 0.063 | 0.002 ± 0.037 | 0.26 | 0.733 |
| L1 | 0.733 ± 0.073 | 0.735 ± 0.075 | 0.001 ± 0.040 | 0.14 | 0.773 |
| L2 | 0.767 ± 0.077 | 0.772 ± 0.074 | 0.006 ± 0.042 | 0.78 | 0.266 |
| L3 | 0.801 ± 0.079 | 0.810 ± 0.075 | 0.009 ± 0.038 | 1.12 | 0.071 |
| L4 | 0.825 ± 0.074 | 0.833 ± 0.067 | 0.008 ± 0.046 | 0.97 | 0.210 |
| *T-score* |  |  |  |  |  |
| Femoral Neck | −1.91 ± 0.63 | −1.92 ± 0.68 | −0.02 ± 0.29 | −1.05 | 0.646 |
| Total Hip | −1.16 ± 0.64 | −1.22 ± 0.66 | −0.05 ± 0.33 | −4.31 | 0.194 |
| Lumbar Spine | −1.77 ± 0.62 | −1.75 ± 0.58 | 0.03 ± 0.32 | 1.69 | 0.495 |
| L1 | −1.76 ± 0.64 | −1.74 ± 0.67 | 0.03 ± 0.39 | 1.70 | 0.594 |
| L2 | −1.88 ± 0.69 | −1.83 ± 0.66 | 0.05 ± 0.37 | 2.66 | 0.281 |
| L3 | −1.84 ± 0.71 | −1.76 ± 0.67 | 0.07 ± 0.35 | 3.80 | 0.110 |
| L4 | −1.81 ± 0.67 | −1.73 ± 0.59 | 0.08 ± 0.41 | 4.42 | 0.182 |

Data are mean ± standard deviation. <sup>a</sup>Paired-T test was conducted. *P*-values < .05 were considered significant.
BMD, bone mineral density.

The time-dependent changes in bone turnover markers over a period of 6 and 12 months are shown in ***Figure 1***. There was no significant change in osteocalcin and the bone resorption markers, NTX and CTX, at the 6-month and 12-month marks (***Figure 1A, 1C*** and ***1D***). Conversely, a significant decrease in the bone formation marker P1NP was observed (***Figure 1B***, both *P* < 0.05).

**Figure 1.**
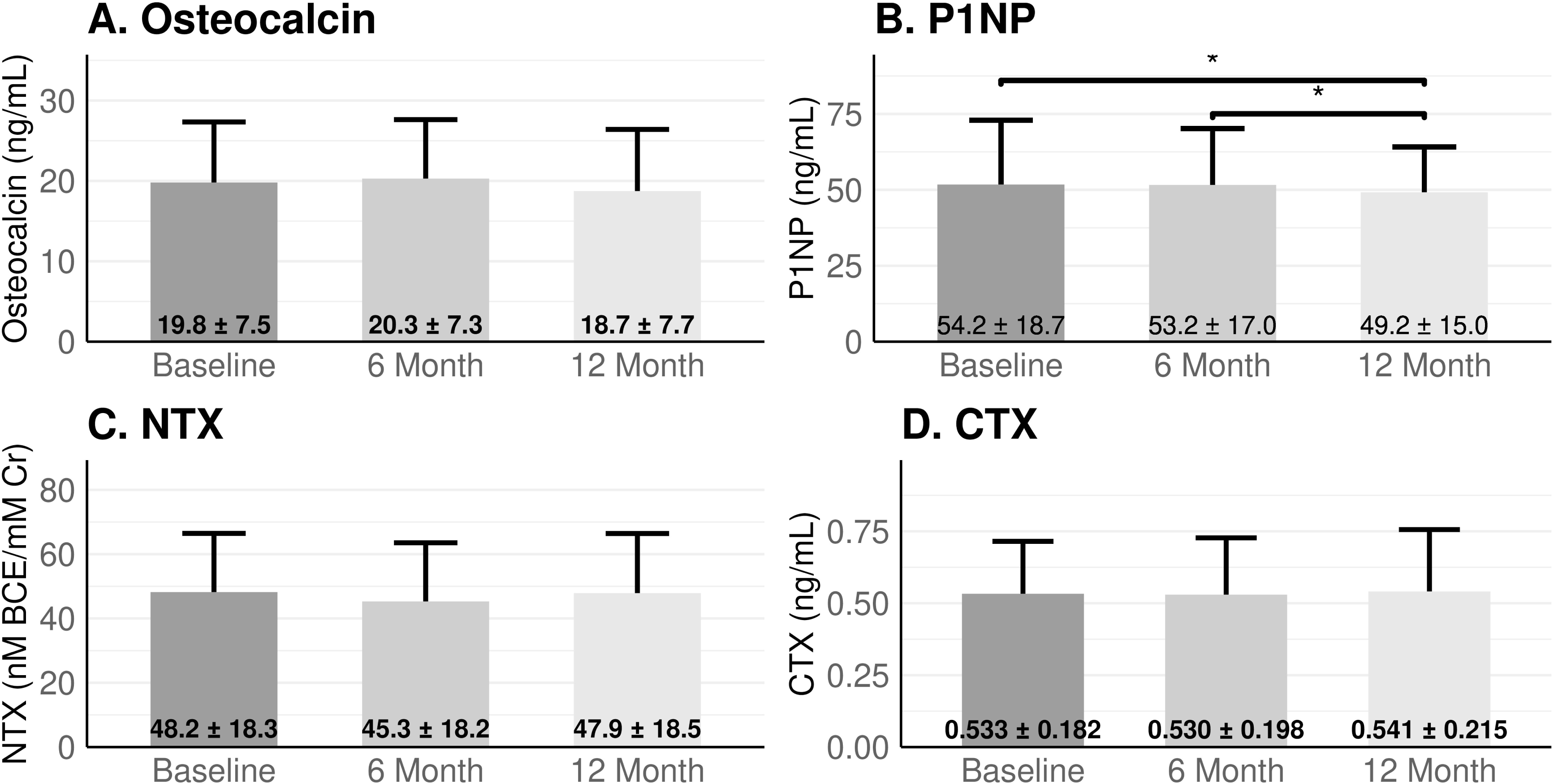
Changes in bone markers at baseline, 6 months, and 12 months. (A) Osteocalcin, (B) P1NP, (C) NTX, and (D) CTX. Data are presented as mean ± SD. A paired-T test indicated a significant difference between the two time points (*P* < 0.05), as marked by an asterisk *. P1NP, procollagen type 1 N-propeptide; NTX, N-terminal telopeptide; CTX, C-terminal telopeptide; SD, standard deviation.

### Extended Observations of the Participants

At the additional follow-up, 63 patients were further evaluated at a median of 26 months (IQR 24.8-31.8). Laboratory values and BMD changes at the third visit are detailed in **Table S2** and **S3**. The decrease in lipid parameters, including total cholesterol, LDL-cholesterol, and triglycerides, was maintained continuously in the extended observation (all *Ps* < 0.001). However, HDL-cholesterol did not change significantly. Additionally, 25OH-vitamin D levels showed a significant increase from baseline to the 26-month follow-up (*P* < 0.001).

In assessing changes in BMD and T-scores over an extended period, we observed significant decreases at the FN: absolute change in BMD was –0.011 ± 0.032 (–0.85%/year, *P* = 0.013), and in the T-score was –0.12 ± 0.36 (–3.01%/year, *P* = 0.011); and at the TH: the absolute change in BMD was –0.027 ± 0.055 (–1.69%/year, *P* < 0.001), and in the T-score was –0.24 ± 0.39 (–10.03%/year, *P* < 0.001). No statistically significant changes were noted at the LS.

Furthermore, we indirectly compared our extended data, which includes examination beyond the second visit year up to four years, with longitudinal studies conducted over 4 years or more on postmenopausal women from five international and two Korean cohorts (**Table S4**). The estimated slope of our cohort, along with the average slopes of international (FN: −0.010, TH: −0.014, LS: −0.021) and Korean (FN: −0.008, TH: −0.008, LS: −0.001) cohorts at each site, are presented in ***Figure 2***. The slopes of the linear fitted unadjusted models for our cohort at the FN (β = –0.003, SE = 0.001, *P* = 0.025), TH (β = –0.005, SE = 0.002, *P* = 0.004), and LS (β = 0.001, SE = 0.002, *P* = 0.862) showed significant differences from the average slopes of the international cohorts (all *Ps* < 0.05). However, compared to the Korean cohorts, the fitted linear slopes for our cohort were not significantly different at the TH (*P* = 0.146) and LS (*P* = 0.937).

**Figure 2.**
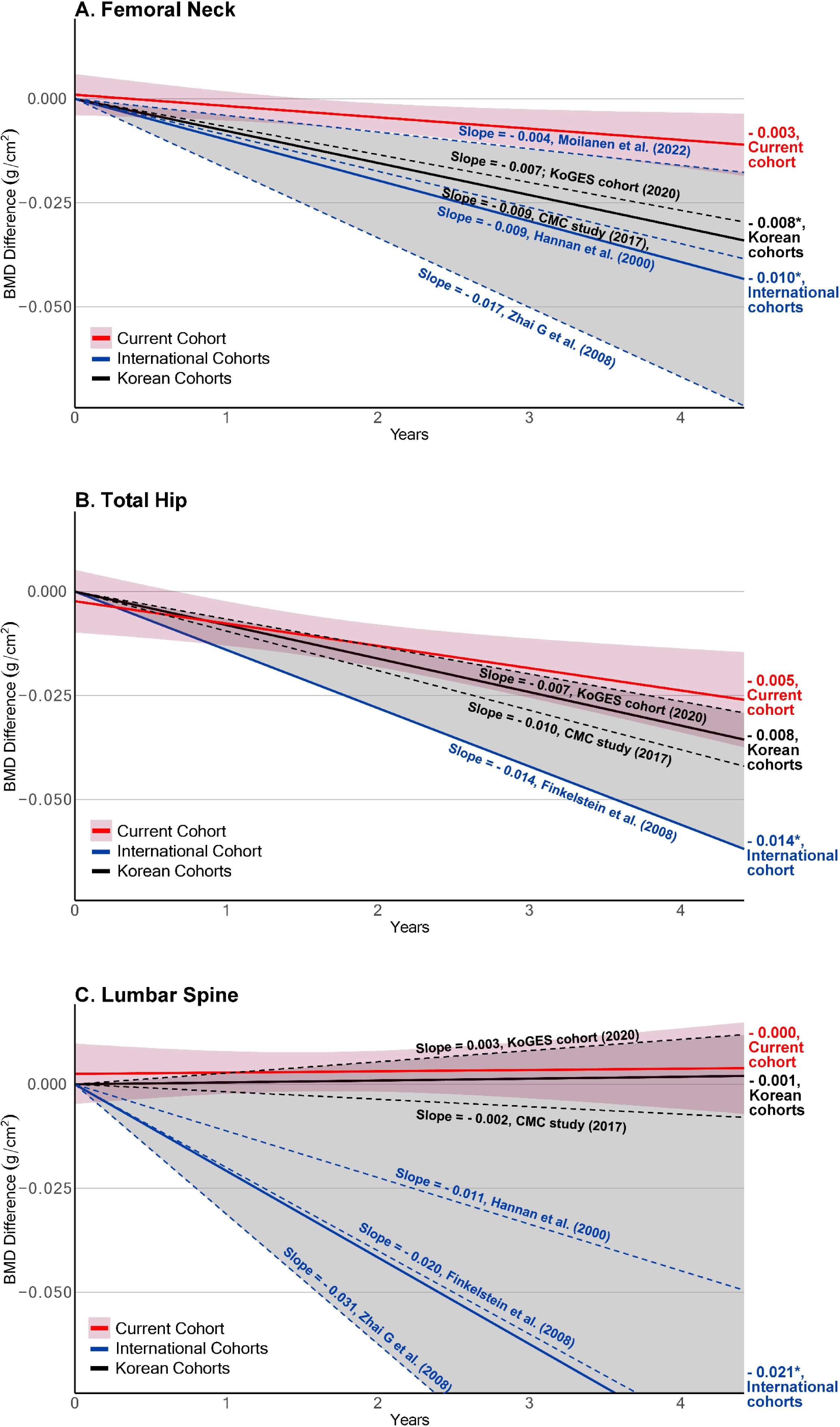
Slope of BMD changes comparing the current cohort using pitavastatin to International and Korean cohorts describing natural decline in menopausal women. (A) Femoral Neck, (B) Total Hip, (C) Lumbar Spine. The figure illustrates the BMD difference (g/cm²) over time (years) for the current cohort (red line), with a red shaded area representing the confidence intervals of the linear fitted model. Participants in this study were observed for up to 4 years (1,614 days), including an examination beyond the second year, by visiting the regular clinic annually. An asterisk * indicates a significant difference between the estimated slope of the current cohort and the averaged reference slopes, represented by solid lines, of the international and Korean cohorts (*P* < 0.05). The individual reference cohort slopes of the international and Korean cohorts, as described in **Table S4**, are shown as dotted lines in blue and black, respectively. The range of slopes for both international and Korean cohorts is represented as a gray shaded area.

### Safety and Tolerability

A total of 29 (41.4%) patients experienced adverse events, most of which were mild severity. The most frequent adverse events (AEs) were musculoskeletal disorders, affecting 13 (17.1%) of patients. All patients continued the medication, with two patients reducing the dosage due to mild myalgia. Eight patients reported gastrointestinal disorders, while three experienced urinary disorders. One spinal fracture was observed as a result of a fell on an icy road, and one serious adverse event, hospitalization due to cholangitis, was observed during the study period. Adverse events summarized by organ in the participants are shown in ***Table 4***.

**Table 4.** Adverse events.

| **Adverse events by organ** | **Patients, n=70** |
| --- | --- |
| Any adverse events - no. (%) | 29 (41.4) |
| Musculoskeletal events - no. (%) | 13 (18.5) |
| Back pain | 2 |
| Knee pain | 2 |
| Muscle pain | 2 |
| Myalgia | 2 |
| Ankle pain | 1 |
| Arthralgia | 1 |
| Chest pain | 1 |
| Cramp in leg | 1 |
| Shoulder pain | 1 |
| Spinal fracture | 1 |
| Tremor | 1 |
| Wrist pain | 1 |
| Gastrointestinal events - no. (%) | 8 (11.4) |
| Dyspepsia | 6 |
| Abdominal pain | 3 |
| Anorexia | 1 |
| Enteritis | 1 |
| Epigastric pain | 1 |
| Nausea | 1 |
| Urinary events - no. (%) | 3 (4.3) |
| Bacteriuria | 1 |
| Flank pain | 1 |
| Irregular bleeding | 1 |
| Other abnormalities- no. (%) | 13 (18.5) |
| Edema | 4 |

Adverse events of different organs were counted independently.

## Discussion

In our study, postmenopausal women with osteopenia or osteoporosis treated with pitavastatin exhibited no significant changes in BMD at FN, TH, and LS sites during the first year. Additionally, pitavastatin therapy significantly reduced LDL-cholesterol levels and LDL subfraction scores, also benefiting cardiovascular risk. The reduction in small, LDL subfraction levels after pitavastatin therapy is a new finding in the current study.^15^ Our findings suggest that pitavastatin is a favorable option for treating dyslipidemia and does not negatively affect BMD or worsen the severity of osteopenia or osteoporosis in postmenopausal women.

Long-term use of pitavastatin therapy was found to be safe in the context of BMD changes compared to the natural decline observed in postmenopausal women (**Table S4** and **Figure S1**).^7,16,17^ After one year of pitavastatin treatment, the percentage changes in BMD were –0.16%/year at the FN, –0.66%/year at the TH, and 0.26%/year at the LS. These changes were similar or less severe than natural declines reported in previous studies.^16–19^ The Framingham Osteoporosis Study, which included postmenopausal women aged 67-95 years, showed declines in BMD of –0.87%/year at the FN during a 4-year follow-up period.^19^ A study in Denmark that investigated BMD changes over a 2-year interval in postmenopausal women aged 60-69 years without conditions affecting bone metabolism, revealed a BMD decrease of –0.40%/2 years in the TH and an increase of 0.50%/2 years in the LS.^18^ Two Korean prospective studies reported annual BMD changes of –0.67% and –0.87% at the FN, –0.66% and 0.95% at the TH, and –0.18% and 0.27% at the LS sites, respectively, among elderly women without osteoporosis treatment.^16^ Compared to these studies, our study showed a smaller decrease at the FN and TH in postmenopausal women with similar age ranges, suggesting pitavastatin may have a neutral or slightly positive effect on bone metabolism.

Preclinical investigations have shown that statins possess anabolic effects on bone by diminishing the synthesis of mevalonate and impairing protein prenylation, thereby stimulating bone morphogenetic protein-2.^20^ However, clinical studies have shown varied effects depending on specific conditions. In a case-control study, in women under 70 years, statin use is associated with decreased osteoporosis rates, likely due to their osteogenic effects, such as promoting osteoblast differentiation and inhibiting osteoclast activation.^21^ Conversely, in women aged 70 years and above, statin use was associated with higher osteoporosis rates, possibly due to the estrogen-lowering effects of statins, which increase bone resorption and reduce bone density.^8,22^

Most studies investigating the effects of statins on BMD had relatively short durations, such as one year. Twelve-month treatment of simvastatin 20 mg did not change BMD at the FN and LS in 32 postmenopausal subjects with an average age of 56.8 years and a BMI of 29.3 ± 4.5 kg/m^2^.^23^ Similarly, in a study involving 318 postmenopausal participants with osteopenia, aged 40-75 years, who were randomly assigned to receive a once-daily dose of atorvastatin (10, 20, 40, or 80 mg) or a placebo, there were no significant changes in BMD at the FN and LS in any treatment group.^24^ Thus, studies investigating the efficacy and safety of statins on bone health have been limited and varied in study design, making it difficult to draw definitive conclusion.

The studies investigating the effects of statins on bone formation and resorption markers have also been limited, yielding mixed results. Osteocalcin levels initially increased and then decreased, indicating the transient effects of statins on bone metabolism. One study reported that a four-week treatment with 20 mg of simvastatin increased osteocalcin levels,^25^ but this positive effect was attenuated with longer treatment durations.^23^ In contrast, our study did not observe any significant changes in osteocalcin levels.

Regarding P1NP, our study showed a significant decrease over 12 months. This finding aligns with a cohort study of 500 statin users (age 67 ± 10 years, BMI 29.7 ± 4.1 kg/m^2^). Among these users, 81 individuals on stable doses of statins (pravastatin, lovastatin, fluvastatin, simvastatin, or atorvastatin) for less than a year exhibited significantly lower P1NP levels compared to 1,931 non-users with similar age and BMI.^26^

In a recent randomized study with 24 postmenopausal women, a statistically significant decrease in NTX was observed in patients receiving rosuvastatin (5 to 10 mg) but not in those receiving atorvastatin 20 mg.^27^ A meta-analysis with 5 studies reported that statin treatment decreased NTX levels (–1.14 nM BCE, 95% CI: –1.21, –0.07).^28^ Experimental studies reported that statins treatment suppressed osteoclast formation through the osteoprotegerin/receptor activator of nuclear factor kappa-B (NFKB) ligand/receptor activator of NFKB.^29,30^ Significantly, our study found no changes in NTX and CTX levels after one year of pitavastatin treatment, indicating that pitavastatin may not increase bone resorption, potentially stabilizing the overall bone turnover rate in postmenopausal women.

In a randomized controlled trial, disparate effects of statins on 25OH-vitamin D levels were reported: over an 8-week treatment period, 10 mg of rosuvastatin increased these levels, whereas 80 mg of fluvastatin did not.^31^ Despite both dosages having equivalent drug potencies, their effects on vitamin D levels differed, suggesting that the impact of statin therapy on vitamin D levels might be inconsistent. The underlying mechanism remains unclear but likely involves the metabolism of vitamin D by CYP3A4 and CYP3A5 enzymes in the liver and intestines—pathways heavily utilized by statins, which could indicate a possible positive influence on vitamin D levels.^32^ Furthermore, in our study, 25OH-vitamin D levels increased significantly over two years. Among these participants, 24 patients (35.3%) regularly took vitamin D and calcium supplements, which remained unchanged during the study period. Although the exact mechanisms by which statins affect vitamin D metabolism are not fully determined,^33^ this could contribute beneficially to statin’s impact on bone health. Taken together, given the variety of factors such as statin type, dosage, treatment duration, and patient characteristics influencing the effects of statins on bone metabolism, well-designed and larger studies are essential to provide definitive results.

In our study, we observed a reduction in LDL-cholesterol levels by 36-39% following the administration of 4 mg of pitavastatin. Furthermore, it decreased non-HDL-cholesterol levels from 167.4 ± 28.2 mg/dL at baseline to 111.6 ± 33.0 mg/dL at the 12-month follow-up. Similarly, in the REAL-CAD study, daily administration of 4 mg of pitavastatin significantly reduced cardiovascular events in 13,054 Japanese patients with stable coronary artery disease (HR = 0.81, 95% CI 0.69-0.95).^34^ A decreased LDL subfraction score due to pitavastatin treatment, as found in our current study, reinforces the effectiveness of reducing cardiovascular risks. An additional benefit of pitavastatin is its demonstrated lower risk of new-onset diabetes mellitus compared to atorvastatin or rosuvastatin.^35,36^

In the current study, a control group was not included. However, establishing a no-medication group could be challenging, as all women with hypercholesterolemia require lipid-lowering medication. Instead, we compared the BMD changes in our study to those observed in population from both Asian as well as Western cohorts with longitudinal observations.

## Conclusions

Our study suggests that pitavastatin therapy can be an ideal option for treating dyslipidemia in postmenopausal women with osteopenia or osteoporosis, without adversely affecting bone health over a period of up to 4 years.

## Affiliations

Department of Internal Medicine, Seoul National University College of Medicine, Seoul National University Bundang Hospital, Seongnam, South Korea (J.H., M.S., H.O., S.L.).

## Supplementary Material

Tables S1-S2

## References

1. Gonyeau MJ. Statins and osteoporosis: a clinical review. Pharmacotherapy. 2005;25:228–243. doi: 10.1592/phco.25.2.228.56954

2. Meier CR, Schlienger RG, Kraenzlin ME, Schlegel B, Jick H. HMG-CoA reductase inhibitors and the risk of fractures. JAMA. 2000;283:3205–3210. doi: 10.1001/jama.283.24.3205

3. LaCroix AZ, Cauley JA, Pettinger M, Hsia J, Bauer DC, McGowan J, Chen Z, Lewis CE, McNeeley SG, Passaro MD, et al. Statin use, clinical fracture, and bone density in postmenopausal women: results from the Women’s Health Initiative Observational Study. Ann Intern Med. 2003;139:97–104. doi: 10.7326/0003-4819-139-2-200307150-00009

4. Wada Y, Nakamura Y, Koshiyama H. Lack of positive correlation between statin use and bone mineral density in Japanese subjects with type 2 diabetes. Arch Intern Med. 2000;160:2865. doi: 10.1001/archinte.160.18.2865

5. Ahn SH, Park SM, Park SY, Yoo JI, Jung HS, Nho JH, Kim SH, Lee YK, Ha YC, Jang S, et al. Osteoporosis and Osteoporotic Fracture Fact Sheet in Korea. J Bone Metab. 2020;27:281–290. doi: 10.11005/jbm.2020.27.4.281

6. Sozen T, Ozisik L, Basaran NC. An overview and management of osteoporosis. Eur J Rheumatol. 2017;4:46–56. doi: 10.5152/eurjrheum.2016.048

7. Finkelstein JS, Brockwell SE, Mehta V, Greendale GA, Sowers MR, Ettinger B, Lo JC, Johnston JM, Cauley JA, Danielson ME, et al. Bone mineral density changes during the menopause transition in a multiethnic cohort of women. J Clin Endocrinol Metab. 2008;93:861–868. doi: 10.1210/jc.2007-1876

8. An T, Hao J, Sun S, Li R, Yang M, Cheng G, Zou M. Efficacy of statins for osteoporosis: a systematic review and meta-analysis. Osteoporos Int. 2017;28:47–57. doi: 10.1007/s00198-016-3844-8

9. Arnett DK, Blumenthal RS, Albert MA, Buroker AB, Goldberger ZD, Hahn EJ, Himmelfarb CD, Khera A, Lloyd-Jones D, McEvoy JW, et al. 2019 ACC/AHA Guideline on the Primary Prevention of Cardiovascular Disease: Executive Summary: A Report of the American College of Cardiology/American Heart Association Task Force on Clinical Practice Guidelines. J Am Coll Cardiol. 2019;74:1376–1414. doi: 10.1016/j.jacc.2019.03.009

10. Mukhtar RY, Reid J, Reckless JP. Pitavastatin. Int J Clin Pract. 2005;59:239–252. doi: 10.1111/j.1742-1241.2005.00461.x

11. Fujino H, Yamada I, Shimada S, Yoneda M, Kojima J. Metabolic fate of pitavastatin, a new inhibitor of HMG-CoA reductase: human UDP-glucuronosyltransferase enzymes involved in lactonization. Xenobiotica. 2003;33:27–41. doi: 10.1080/0049825021000017957

12. Majima T, Shimatsu A, Komatsu Y, Satoh N, Fukao A, Ninomiya K, Matsumura T, Nakao K. Short-term effects of pitavastatin on biochemical markers of bone turnover in patients with hypercholesterolemia. Intern Med. 2007;46:1967–1973. doi: 10.2169/internalmedicine.46.0419

13. Diab DL, Watts NB. Diagnosis and treatment of osteoporosis in older adults. Endocrinol Metab Clin North Am. 2013;42:305–317. doi: 10.1016/j.ecl.2013.02.007

14. Hoefner DM, Hodel SD, O’Brien JF, Branum EL, Sun D, Meissner I, McConnell JP. Development of a rapid, quantitative method for LDL subfractionation with use of the Quantimetrix Lipoprint LDL System. Clin Chem. 2001;47:266–274.

15. Choi CU, Seo HS, Lee EM, Shin SY, Choi UJ, Na JO, Lim HE, Kim JW, Kim EJ, Rha SW, et al. Statins do not decrease small, dense low-density lipoprotein. Tex Heart Inst J. 2010;37:421–428.

16. Park SY, Kim JH, Choi HJ, Ku EJ, Hong AR, Lee JH, Shin CS, Cho NH. Longitudinal changes in bone mineral density and trabecular bone score in Korean adults: a community-based prospective study. Arch Osteoporos. 2020;15:100. doi: 10.1007/s11657-020-00731-6

17. Lim Y, Jo K, Ha HS, Yim HW, Yoon KH, Lee WC, Son HY, Baek KH, Kang MI. The prevalence of osteoporosis and the rate of bone loss in Korean adults: the Chungju metabolic disease cohort (CMC) study. Osteoporos Int. 2017;28:1453–1459. doi: 10.1007/s00198-016-3893-z

18. Warming L, Hassager C, Christiansen C. Changes in bone mineral density with age in men and women: a longitudinal study. Osteoporos Int. 2002;13:105–112. doi: 10.1007/s001980200001

19. Hannan MT, Felson DT, Dawson-Hughes B, Tucker KL, Cupples LA, Wilson PW, Kiel DP. Risk factors for longitudinal bone loss in elderly men and women: the Framingham Osteoporosis Study. J Bone Miner Res. 2000;15:710–720. doi: 10.1359/jbmr.2000.15.4.710

20. Zhang Y, Bradley AD, Wang D, Reinhardt RA. Statins, bone metabolism and treatment of bone catabolic diseases. Pharmacol Res. 2014;88:53–61. doi: 10.1016/j.phrs.2013.12.009

21. Kim SY, Yoo DM, Min C, Kim JH, Kwon MJ, Kim JH, Choi HG. Association between Osteoporosis and Previous Statin Use: A Nested Case-Control Study. Int J Environ Res Public Health. 2021;18. doi: 10.3390/ijerph182211902

22. Bone HG, Greenspan SL, McKeever C, Bell N, Davidson M, Downs RW, Emkey R, Meunier PJ, Miller SS, Mulloy AL, et al. Alendronate and estrogen effects in postmenopausal women with low bone mineral density. Alendronate/Estrogen Study Group. J Clin Endocrinol Metab. 2000;85:720–726. doi: 10.1210/jcem.85.2.6393

23. Tikiz C, Tikiz H, Taneli F, Gumuser G, Tuzun C. Effects of simvastatin on bone mineral density and remodeling parameters in postmenopausal osteopenic subjects: 1-year follow-up study. Clin Rheumatol. 2005;24:447–452. doi: 10.1007/s10067-004-1053-x

24. Bone HG, Kiel DP, Lindsay RS, Lewiecki EM, Bolognese MA, Leary ET, Lowe W, McClung MR. Effects of atorvastatin on bone in postmenopausal women with dyslipidemia: a double-blind, placebo-controlled, dose-ranging trial. J Clin Endocrinol Metab. 2007;92:4671–4677. doi: 10.1210/jc.2006-1909

25. Chan MH, Mak TW, Chiu RW, Chow CC, Chan IH, Lam CW. Simvastatin increases serum osteocalcin concentration in patients treated for hypercholesterolaemia. J Clin Endocrinol Metab. 2001;86:4556–4559. doi: 10.1210/jcem.86.9.8001

26. Hernandez JL, Olmos JM, Romana G, Martinez J, Castillo J, Yezerska I, Ramos C, Gonzalez-Macias J. Bone turnover markers in statin users: a population-based analysis from the Camargo Cohort Study. Maturitas. 2013;75:67–73. doi: 10.1016/j.maturitas.2013.02.003

27. Braszak-Cymerman A, Walczak MK, Oduah MT, Ludziejewska A, Bryl W. Comparison of the pleiotropic effect of atorvastatin and rosuvastatin on postmenopausal changes in bone turnover: A randomized comparative study. Medicine (Baltimore). 2024;103:e38122. doi: 10.1097/MD.0000000000038122

28. Zhao H, Tang Y, Zhen Y, Qi C, Chen S. The effect of statins on bone turnover biomarkers: a systematic review and meta-analysis of randomized controlled trials. Endocr J. 2023;70:473–480. doi: 10.1507/endocrj.EJ22-0512

29. Sun X, Wei B, Peng Z, Fu Q, Wang C, Zhen J, Sun J. Protective effects of Dipsacus asper polysaccharide on osteoporosis in vivo by regulating RANKL/RANK/OPG/VEGF and PI3K/Akt/eNOS pathway. Int J Biol Macromol. 2019;129:579–587. doi: 10.1016/j.ijbiomac.2019.02.022

30. Elewa HF, El-Remessy AB, Somanath PR, Fagan SC. Diverse effects of statins on angiogenesis: new therapeutic avenues. Pharmacotherapy. 2010;30:169–176. doi: 10.1592/phco.30.2.169

31. Ertugrul DT, Yavuz B, Cil H, Ata N, Akin KO, Kucukazman M, Yalcin AA, Dal K, Yavuz BB, Tutal E. STATIN-D study: comparison of the influences of rosuvastatin and fluvastatin treatment on the levels of 25 hydroxyvitamin D. Cardiovasc Ther. 2011;29:146–152. doi: 10.1111/j.1755-5922.2010.00141.x

32. Xu Y, Hashizume T, Shuhart MC, Davis CL, Nelson WL, Sakaki T, Kalhorn TF, Watkins PB, Schuetz EG, Thummel KE. Intestinal and hepatic CYP3A4 catalyze hydroxylation of 1alpha,25-dihydroxyvitamin D(3): implications for drug-induced osteomalacia. Mol Pharmacol. 2006;69:56–65. doi: 10.1124/mol.105.017392

33. Mazidi M, Rezaie P, Vatanparast H, Kengne AP. Effect of statins on serum vitamin D concentrations: a systematic review and meta-analysis. Eur J Clin Invest. 2017;47:93–101. doi: 10.1111/eci.12698

34. Taguchi I, Iimuro S, Iwata H, Takashima H, Abe M, Amiya E, Ogawa T, Ozaki Y, Sakuma I, Nakagawa Y, et al. High-Dose Versus Low-Dose Pitavastatin in Japanese Patients With Stable Coronary Artery Disease (REAL-CAD): A Randomized Superiority Trial. Circulation. 2018;137:1997–2009. doi: 10.1161/CIRCULATIONAHA.117.032615

35. Choi JY, Choi CU, Hwang SY, Choi BG, Jang WY, Kim DY, Kim W, Park EJ, Lee S, Na JO, et al. Effect of Pitavastatin Compared with Atorvastatin andRosuvastatin on New-Onset Diabetes Mellitus in PatientsWith Acute Myocardial Infarction. Am J Cardiol. 2018;122:922–928. doi: 10.1016/j.amjcard.2018.06.017

36. Seo WW, Seo SI, Kim Y, Yoo JJ, Shin WG, Kim J, You SC, Park RW, Park YM, Kim KJ, et al. Impact of pitavastatin on new-onset diabetes mellitus compared to atorvastatin and rosuvastatin: a distributed network analysis of 10 real-world databases. Cardiovasc Diabetol. 2022;21:82. doi: 10.1186/s12933-022-01524-6

